# The Effect of Mindfulness Based Stress Reduction on Glycemic Control and Psychological Well-being of Diabetes Mellitus Patients: A Review of Experimental Studies

**DOI:** 10.1101/2023.12.27.23300572

**Authors:** Arina Qona’ah, Moses Glorino Rumambo Pandin, Nursalam Nursalam

## Abstract

Most diabetes mellitus patients experience psychological problems resulting from the disease, disease management, and complications. Inadequate management of psychological problems may place patients at greater risk for depression. This systematic review aims to identify the effects of mindful interventions in controlling glycemic levels and psychological well-being in diabetes mellitus patients. The systematic review conducted on the study used the Preferred Reporting Items for Systematic Review and Meta-analysis (PRISMA) guidelines. This systematic review uses the Scopus, Web of Science, Science Direct, and Google Scholar databases. The keywords used are diabetes mellitus type 2, mindfulness-based stress reduction, AND/OR, glycemic control, blood glucose, glycated hemoglobin, and psychological well-being. The words used are adjusted to the Medical Subject Heading (MeSH). The articles used are published in English in 2014 – 2023, full-text, and are research articles. The search results showed that most mindfulness interventions used were MBSR (Mindfulness-Based Stress Reduction) for 8 weeks taught by professionals. Mindfulness is provided by trained professionals and taught again using communication media, namely CDs or mobile applications. Mindfulness has a significant effect on reducing fasting blood sugar levels and HbA1c in type 2 DM patients. Mindfulness improves psychological well-being and reduces depression, anxiety, resilience, and emotional health in diabetes patients. Mindfulness is a therapy that can be used to manage the psychological problems of DM patients where with good emotional control the patient can control glycemic levels to normal limits

## Background

Diabetes Mellitus (DM) is a chronic disease caused by chronic endocrine metabolic disorders which cause disruption of insulin secretion and/or action, resulting in increased blood sugar levels (hyperglycemia) (Zhang et al., 2022). Diabetes is a complex and debilitating disease that, if not properly controlled, will have negative health impacts (McCoy & Theeke, 2019). The main psychosocial problems faced by DM patients are stress, depression or diabetes distress (Mwila et al., 2019). Nearly 66% of type 2 DM patients experience psychological problems due to diabetes and are at greater risk of developing depression (Watson et al., 2015). These psychological problems negatively impact blood glucose levels, treatment, and disease progression (McCoy & Theeke, 2019).

In 2023, WHO said that the number of DM patients in the world will reach 422 million people. Based on data from *the International Diabetes Federation* in 2021, Indonesia is the fifth DM ranked country with the number of DM sufferers at 19.5 million and it is estimated that this will increase to 28.6 million in 2045 (International Diabetes Federation, 2021). In 2020, DM was the top ten disease with a total of 21,992 people (Dinkes Lamongan, 2021). The results of examination of HbA1c levels in DM patients at the Sukodadi Lamongan Community Health Center in February 2023 were 8.8%, which means the patient’s ability to control blood sugar levels is still low. From the results of research by Mohamed et al in 2023, it was stated that only 5% of DM patients were able to control glucose very good (El-Radad et al., 2023).

Self-management of type 2 DM patients has many aspects including education, treatment, and lifestyle changes. The challenges and problems faced by DM patients are changes in diet and activity habits after being diagnosed with the disease, awareness of the understanding of the disease, and lack of social support (Al Slamah et al., 2020). Barriers to self-management include educational materials, physical, psychological and social barriers, strict treatment regimens, low motivation, lack of understanding of self-management, lack of understanding of health workers towards patients (Chepulis et al., 2021). Failure in self-management can trigger diabetes distress in DM patients (Shahady & O’Grady, 2015).

Diabetes distress is worry and negative emotions related to the experience of living as a diabetic, self-management and complications of the disease (De Wit et al., 2022). The prevalence of diabetes distress is higher in patients with female gender, younger age, shorter duration of diabetes, high HbA1c levels, insulin use, smoking, difficulty following dietary and medication recommendations, insufficient physical activity, and inability to monitor glucose levels. blood (Nanayakkara et al., 2018). Distress causes a narrowed attention span, limited creative solutions, and poor coping and self-management (Guo et al., 2019). Distress can become an obstacle in acquiring new knowledge and skills and the ability to overcome problems. Distress also causes patients to develop unrealistic goals and expectations, inaccurate personal beliefs, and self-defeating perceptions (Shahady & O’Grady, 2015).

One of the strategies that can be used to overcome psychological problems experienced by diabetes patients is mindfulness. MBSR is a series of mindfulness practices that are used to train attention control over current conditions without being accompanied by a judgmental attitude (Crane et al., 2017). MBSR has positive effects on pain, anxiety and stress in people with chronic disorders, such as fibromyalgia, coronary artery disease, back pain and arthritis (Rosenzweig et al., 2010). Systematic reviews show that low to moderate doses of MBSR impact the control of psychological and physical symptoms in a variety of chronic somatic conditions including cancer, cardiovascular disorders, and arthritis (Abbott et al., 2014) (Bohlmeijer et al., 2010). This review aims to identify the effects of mindfulness interventions in controlling glycemic levels and improving psychological well-being in diabetes mellitus patients.

## METHOD

The systematic review conducted on the study used the *Preferred Reporting Items for Systematic Review and Meta-analysis* (PRISMA) guidelines (Shamseer et al., 2015).

Inclusion criteria used PICOS which included patients with type 2 diabetes mellitus (Population); mindfulness intervention (Intervention); a group that refers to a control group or comparison group that does not receive the intervention of music, visual arts, and a combination of both (Control); research includes fasting blood sugar levels, HbA1c, stress, psychological distress, psychological well-being (O); experimental type quantitative research (S); English language articles from 2014 to 2023. Exclusion criteria include articles with the type study protocols, conference presentations, editorials, review articles, case reports, case series, qualitative research.

### Search Strategy

Scannig of academic databases was carried out November 2023. The search was carried out on four databases (Scopus, Web of Science, Science Direct) and we added articles with manual search strategies and direct searches from Google Scholar. The literature search was carried out using Boolean operators in the form of AND OR, namely "diabetes mellitus type 2", " mindfulness-based stress reduction", "AND/OR", "glycemic control", "blood glucose", " Glycated Hemoglobin ", and " psychological well-being". The words used are adjusted to *the Medical Subject Heading* (MeSH).

### Study Selection

Articles were selected using Mendeley. The remaining articles will be assessed and screened by researchers in two different stages namely title and abstract screening, followed by full text screening. Then, the authors independently reviewed the full text. In the screening process, all voting must be blind, meaning my peers cannot see my vote until they cast their own votes, and vice versa. Subsequently, conflicts or disagreements between reviewers were resolved by a third reviewer or consensus-based discussion.

In the study selection process, researchers use the PRISMA Flowchart visual flow chart which assists researchers in identifying, selecting, and organizing studies to be included in a systematic review or meta-analysis. The PRISMA Flowchart is designed to ensure that the study selection process is transparent and replicable (Figure 1).

**Figure 1.**
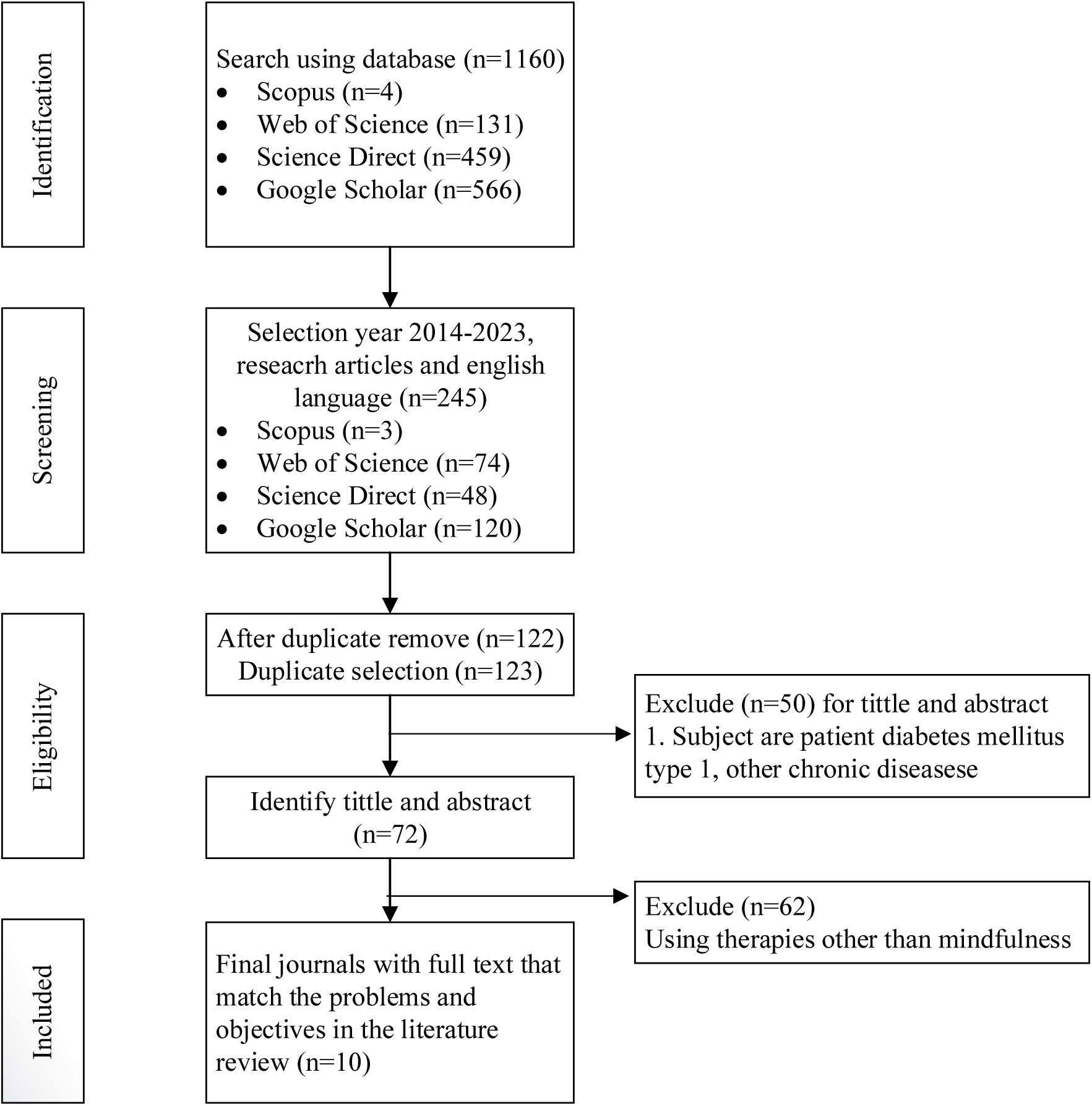
Flowchart with Prisma Guidelines in the Research The Effect of Mindfulness Based Stress Reduction on Glycemic Control and Psychological Well-being of Diabetes Mellitus Patients: A Review of Experimental Studies

### Data Extraction

The inclusion criteria established in each paper were extracted using a Microsoft Excel sheet. Data extraction was carried out independently by two independent researchers. The extraction process must be carried out by a panel of 2-3 impartial reviewers.

### Data analysis

The search flow and literature findings are presented in the form of text and PRISMA-based diagrams. A summary of the evidence that will be used in data synthesis is presented in the form of a summary containing the characteristics of each study. Descriptive data is presented in text and table form.

## RESULTS

### Study Characteristics

Some of the midnfulness research was conducted in the Asian countries including Iran (Kian et al., 2018) (Hosseini et al., 2021)(Ravari et al., 2020), Saudi Arabia (Obaya et al., 2023) and China (Guo et al., 2022). Other countries that have conducted studies on mindfulness are Australia (Pearson et al., 2018), Canada (Dreger et al., 2015)and the United States (DiNardo et al., 2022) (Xia et al., 2022) (Whitebird et al., 2018). In conducting mindfulness research, most researchers use two groups, namely a treatment group (mindfulness or a combination with other therapies) and a control group in the form of education, regular care or physical activity (Kian et al., 2018) (Hosseini et al., 2021) (Pearson et al., 2018) (Ravari et al., 2020) (DiNardo et al., 2022) (Obaya et al., 2023) (Guo et al., 2022). The research was conducted on type 2 DM patients with an average age of 45.24 – 63.7. The respondents involved in this research were mostly male and female except for research (Obaya et al., 2023) (Ravari et al., 2020) which only used women as respondents. The research was conducted on type 2 DM patients who had previously been screened for psychological problems (diabetes distress, stress) using a questionnaire with HbA1C levels > 7, suffering from diabetes for at least six months. The research was not conducted on respondents who were medically diagnosed as suffering from mental problems (Table 1).

**Table 1.**
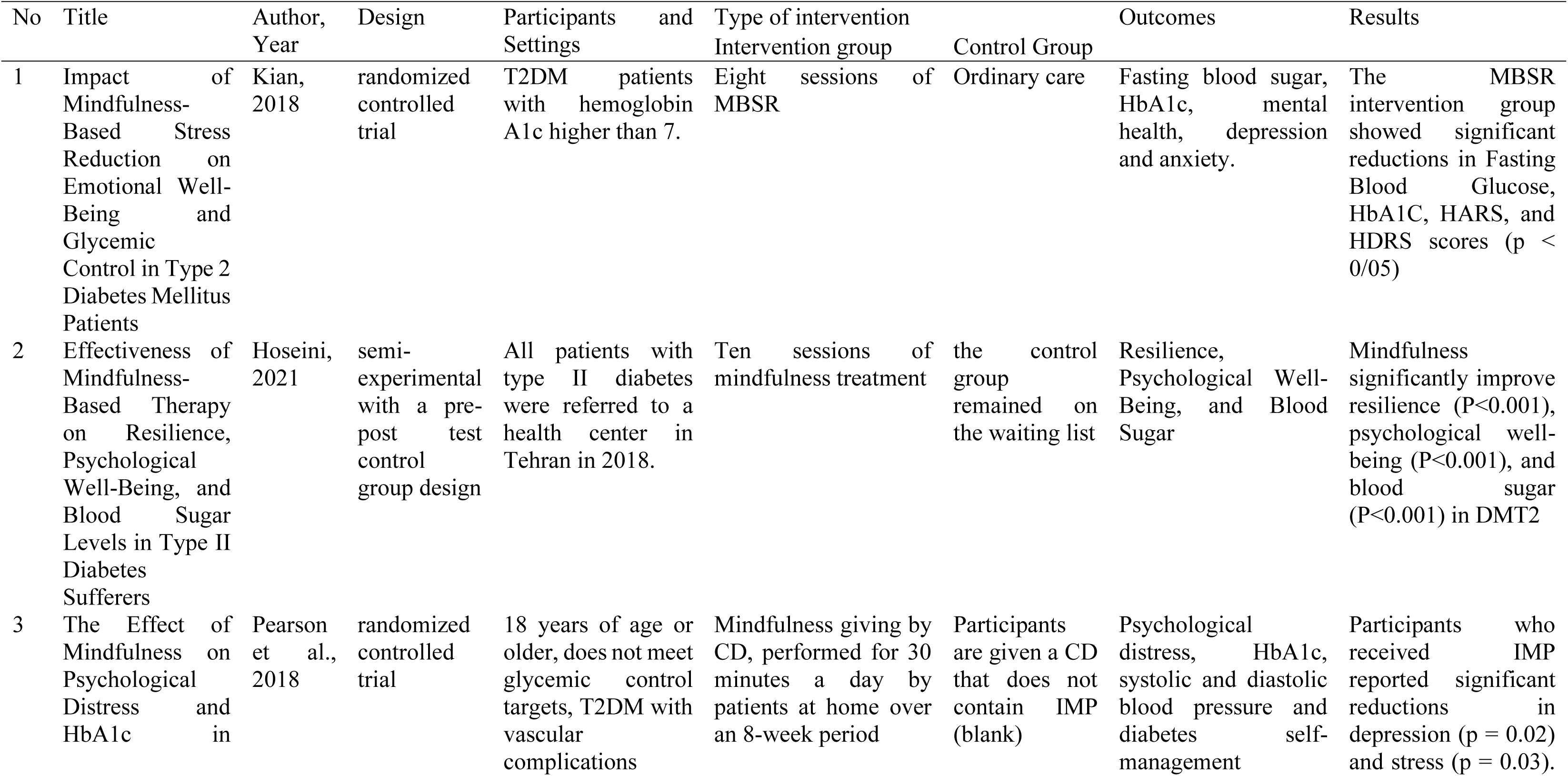
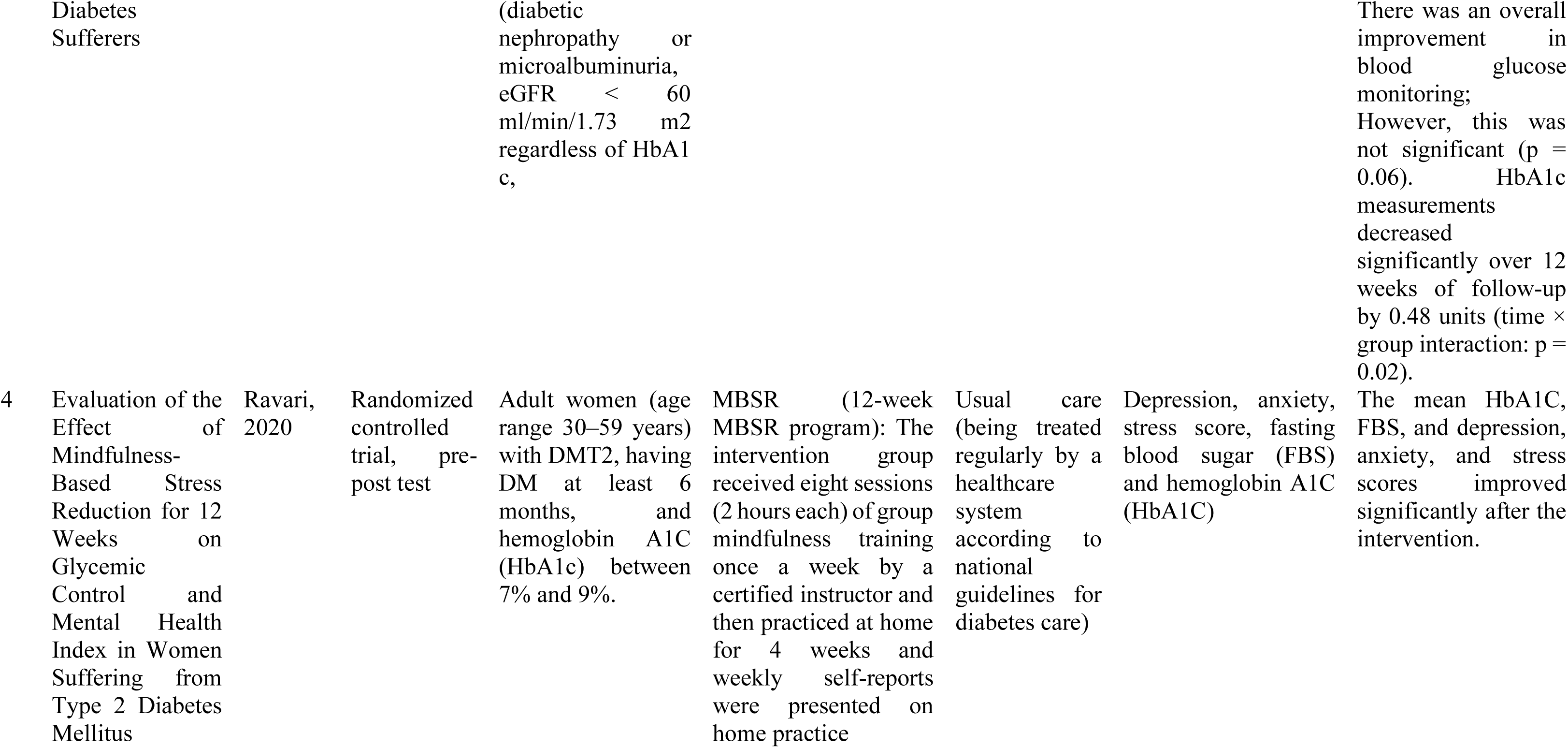
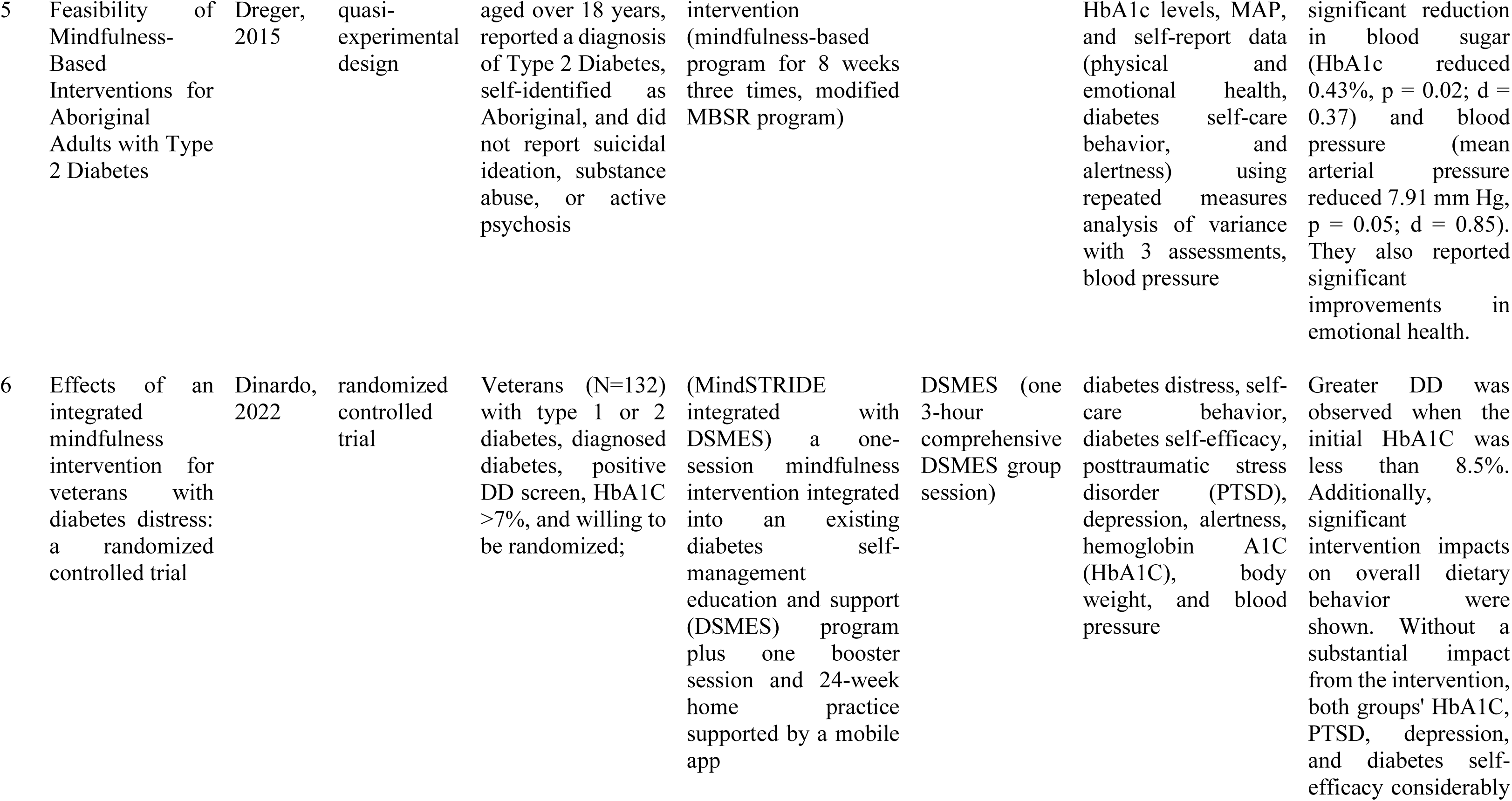
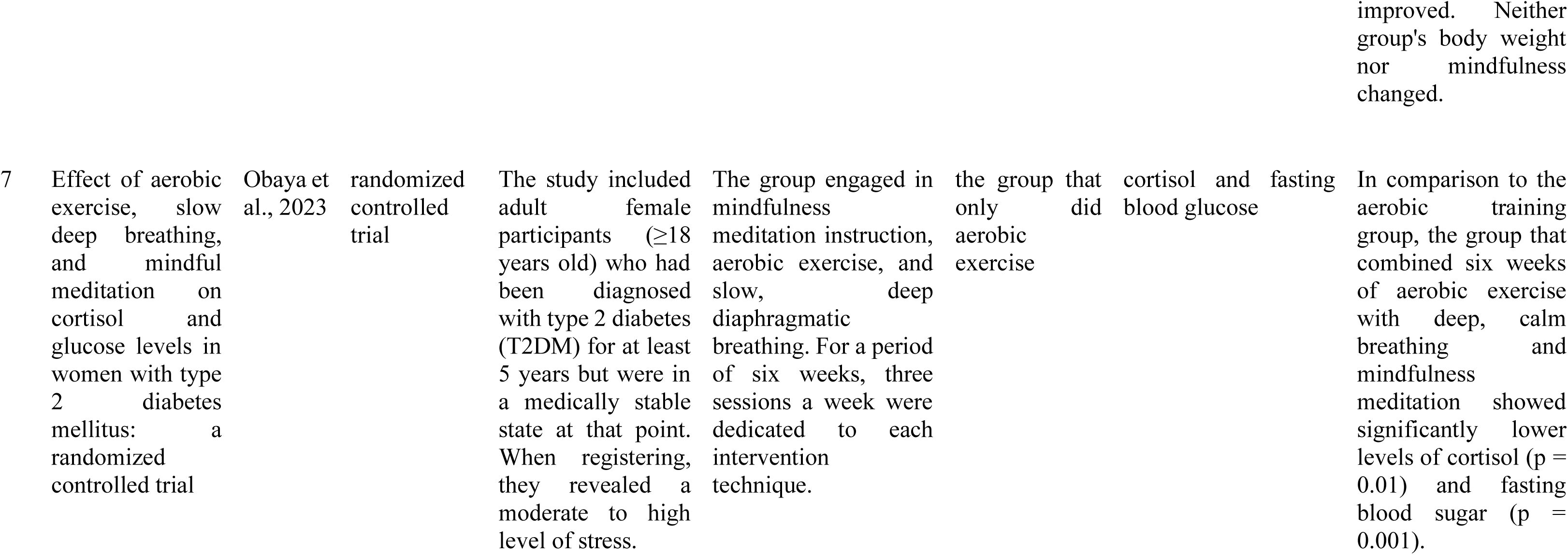
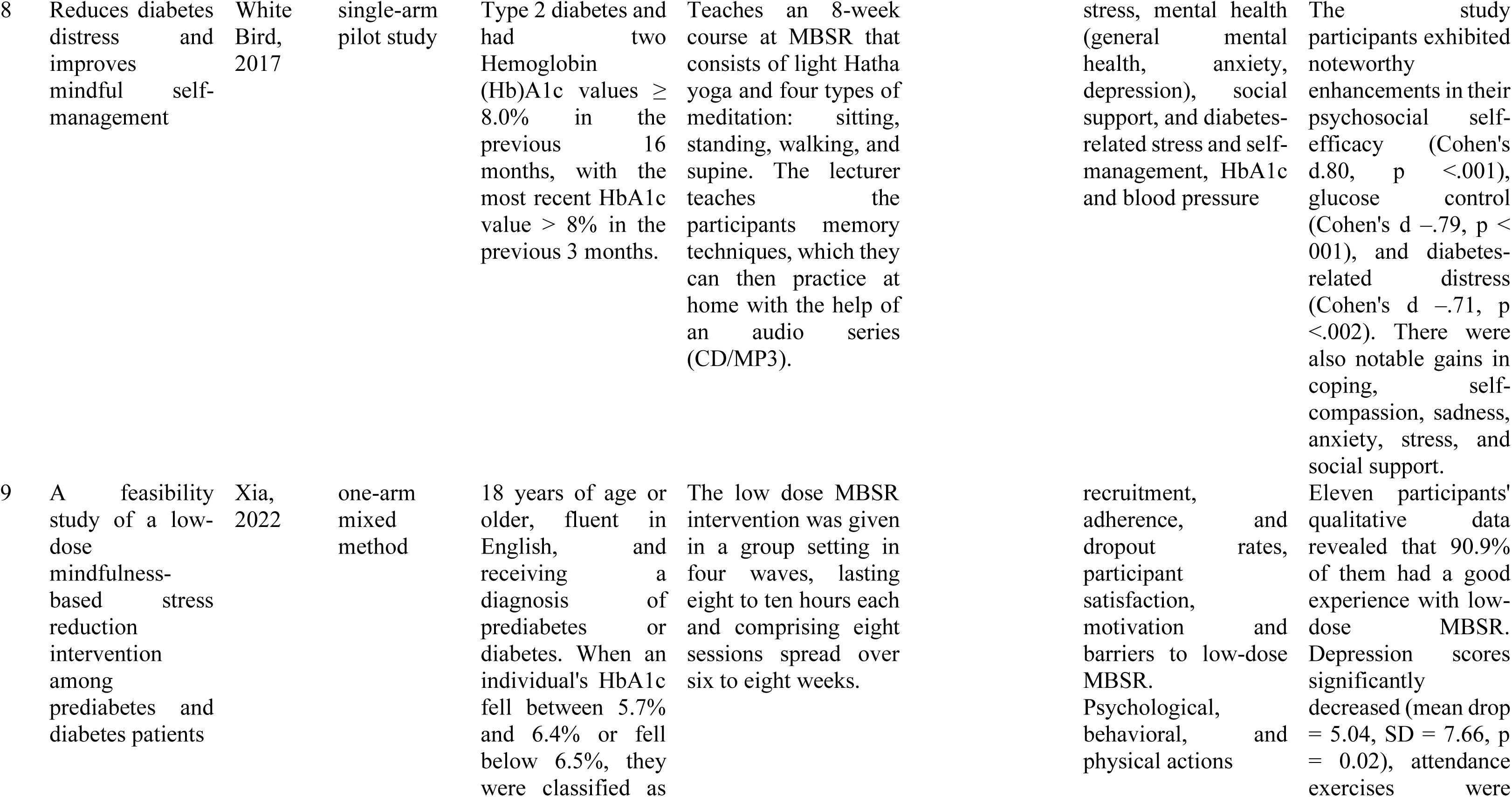
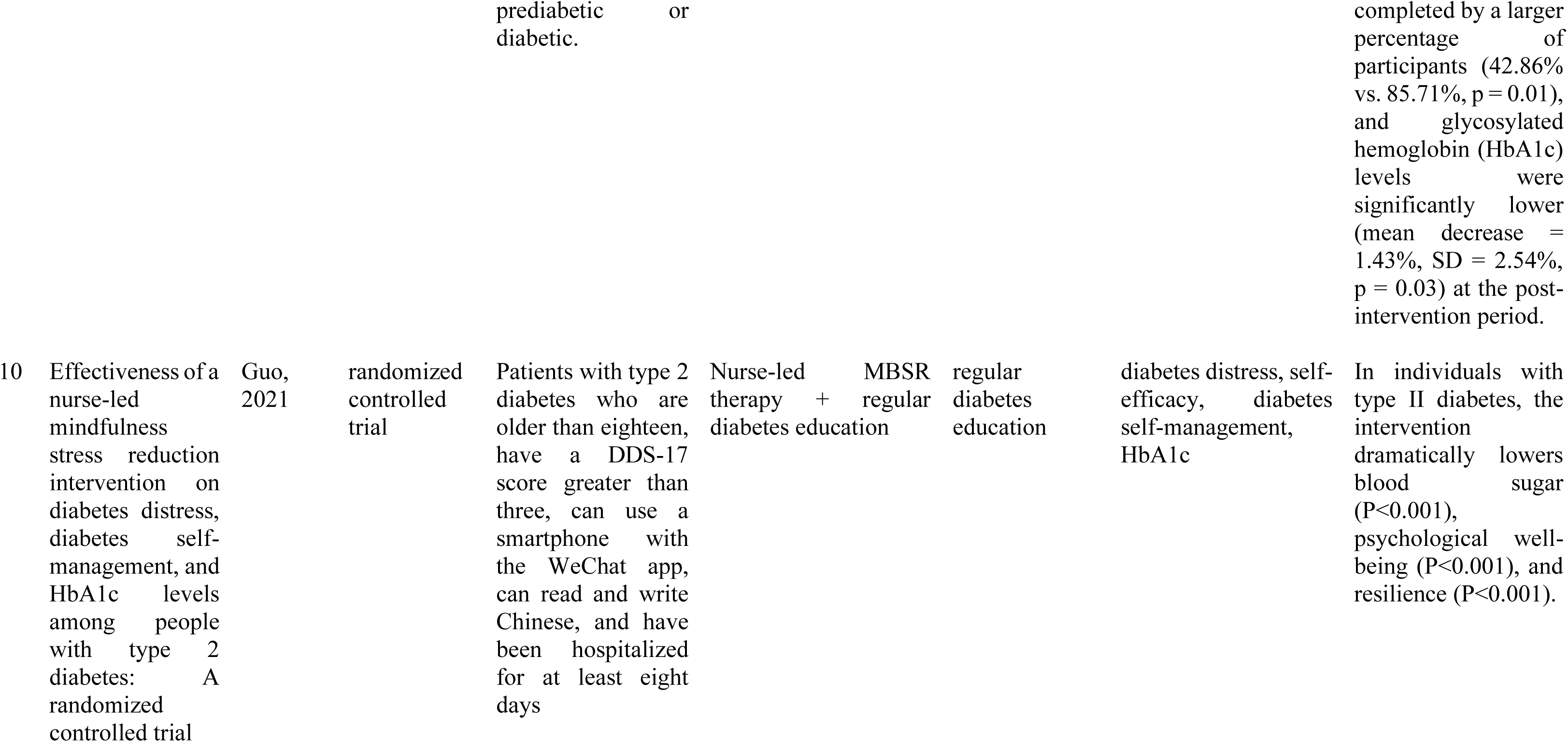
Results of Literature Analysis.

### Mindfulness Intervention

The type of mindfulness intervention used in most research is MBSR (Mindfulness Based Stress Reduction) (Kian et al., 2018) (Ravari et al., 2020) (Dreger et al., 2015) (DiNardo et al., 2022) (Whitebird et al., 2018) (Xia et al., 2022) (Guo et al., 2022). MBSR is carried out for 4 weeks (low dose) (Xia et al., 2022), 8 weeks (Dreger et al., 2015) (Kian et al., 2018) (Pearson et al., 2018) (Dreger et al., 2015) (DiNardo et al., 2022) (Whitebird et al., 2018) and 12 weeks (Ravari et al., 2020). Mindfulness is provided by trained professionals and taught again using communication media, namely CDs (Pearson et al., 2018) (Whitebird et al., 2018) or mobile application (DiNardo et al., 2022).

### Glycemic Control in Type 2 DM Patients Treated with Mindfulness

The effectiveness of mindfulness in controlling glycemic levels in Type 2 DM patients was assessed using fasting blood sugar levels and HbA1c. Measurements were carried out before the intervention, in the middle of the intervention, at the end of the intervention and several months post-intervention (3 months post-intervention) (Kian et al., 2018) (6 months post-intervention) (DiNardo et al., 2022). The research results showed a significant effect of mindfulness in controlling blood sugar and HbA1c levels in type 2 DM patients except in the study (Pearson et al., 2018) where the HbA1c value was significant after measurements were taken at week 12.

### The Effect of Mindfulness on Psychological Well-being

Mindfulness has an effect on controlling the patient’s psychological problems, especially those related to inadequate self-management. MBSR programs increase the wellbeing in type 2 DM patients (Kian et al., 2018)namely by reducing depression (DiNardo et al., 2022) (Xia et al., 2022), depression and anxiety (HARS, and HDRS scores with p < 0.05), reducing depression and stress (Pearson et al., 2018), improvement of resilience and psychological well-being (Hosseini et al., 2021), (Guo et al., 2022) emotional health (Dreger et al., 2015). Apart from being assessed using instruments to evaluate psychological conditions, psychological effects are also assessed from the results of laboratory values, namely cortisol. In research (Obaya et al., 2023) it was found that mindfulness had a significant effect in controlling cortisol values in women.

## DISCUSSION

Mindfulness is an awareness that arises from paying attention deliberately, in the moment, and non-judgmentally to the unfolding of experience moment by moment. Mindfulness refers to a person’s capacity to pay attention to current experiences without judgment or evaluation (Norouzi et al., 2020). Mindfulness Based Intervention was developed for people with chronic physical problems, managing pain, low mood and health-related anxiety (Alsubaie et al., 2017). Mindfulness Based Stress Reduction (MBSR) is a structured program that combines meditation techniques, body awareness, non-judgmental acceptance, and emotional regulation strategies aimed at helping overcome emotional problems. A standard MBSR intervention includes eight weekly group sessions (usually 2.5 hours per session) for 8 weeks with an all-day retreat between weeks 6–7 (Xia et al., 2022). Mindfulness training seeks to increase acceptance of different states of consciousness through a specific focus on physical and emotional discomfort and teaches clients to observe emotional, physical, and cognitive states without involuntary reactions (Hosseini et al., 2021).

Mindfulness is awareness that is non-judgmental and cannot be expressed in words. It is based on an individual’s experience within the limits of his or her attention at a particular moment. In addition, this concept includes acceptance of these experiences and increased mindfulness in an effort to improve psychological well-being. This is as explained in Kian and Hosseini’s research (Kian et al., 2018)(Hosseini et al., 2021) where mindfulness therapy is effective in improving psychological well-being. The components of acceptance, understanding and personal growth are effective, allowing the individual to respond inevitably and contemplatively to events without any contemplation and thought. Additionally, they help people in recognizing, managing, and solving their daily problems. Several studies have shown the effect of increasing mindfulness on improving psychological well-being, life satisfaction, self-esteem, optimism, as well as reducing anxiety, depression, and psychological symptoms (Zimmermann et al., 2018).

Mindfulness can help change behavior especially if targeted at specific behaviors such as diet, physical activity, glucose monitoring, and medication management. In some studies, specifically targeted interventions such as eating control can reduce energy and calorie intake (Miller et al., 2014). Mindfulness training may aid glucose regulation through its effects on emotional regulation and stress reduction. Potential mechanisms or pathways that may explain this HbA1c shift include behavioral changes and stress-reducing effects on the hypothalamic-pituitaryadrenal (HPA) axis through modulation of the HPA axis and stress pathways. In research (Obaya et al., 2023) it was found that mindfulness training reduced cortisol levels in DM patients. The results of research conducted by Rosenzweig et al. showed a decrease in HbA1c levels and an increase in the patient’s ability to manage stress, manage emotional stress, and quality of life. Other research also produces the same thing, namely that mindfulness can improve the psychological coping skills of diabetes patients (Hartmann et al., 2012) (Teixeira, 2010).

## CONCLUSION

Mindfulness Based Stress Reduction (MBSR) is a series of mindfulness practices that are used to train attention control over current conditions without being accompanied by a judgmental attitude. Mindfulness therapy has been proven to be effective in controlling glycemic levels as assessed by fasting blood sugar levels and HbA1c and is able to overcome patients’ psychological problems (proven to be effective in controlling emotions and improving the psychological well-being of diabetes patients). Mindfulness can be a patient companion therapy as an effort to improve self-management efforts for diabetes mellitus patients.

## Data Availability

All data produced in the present study are available upon reasonable request to the authors

